# Barriers and Facilitators of appropriate Antibiotic Dispensing Practices Among Community Settings in Ghana and the implications: A qualitative study

**DOI:** 10.1101/2025.11.24.25340917

**Authors:** George Akafity, Joseph Elikem Efui Acolatse, Marta Wanat, Robert Incoom, Audu Rauf, R Andrew Seaton, Jacqueline Sneddon, Elaine Cameron, Eric Kofi Ngyedu, Brian Godman, Amanj Kurdi

**Author notes:** **JOINT CORRESPONDING AUTHORS**: Professor Amanj Kurdi Strathclyde Institute of Pharmacy and Biomedical Science University of Strathclyde, Glasgow 161 Cathedral Street, G4 0RE. George Akafity Research Monitoring and Evaluation Unit Cape Coast Teaching Hospital CC-070-9967.

## Abstract

Antimicrobial resistance (AMR) remains a critical global health concern, particularly in low- and middle-income countries (LMICs), where inappropriate antibiotic use is widespread. In Ghana, antibiotics are frequently dispensed without prescriptions by community pharmacies (CPs) and over-the-counter medicine sellers (OTCMS), despite regulatory restrictions. This study aimed to explore the behavioural and contextual factors influencing antibiotic dispensing practices among CP and OTCMS personnel using behavioural science frameworks. A qualitative study was conducted using semi-structured interviews with 12 participants across four Ghanaian regions, including eight from CPs (five pharmacists, two pharmacy technicians, and one medical counter assistant) and four from OTCMS outlets. Participants were purposively selected to reflect variation in practice type, setting, and antibiotic dispensing behaviour. Data were thematically analysed and mapped onto the Capability, Opportunity, Motivation-Behavior (COM-B) model and the Theoretical Domains Framework (TDF). Three major themes were identified: perceived readiness and capability to dispense appropriately, the impact of systemic and contextual factors, and motivation to adhere to professional guidelines. Key barriers included limited AMR knowledge among OTCMS staff, weak regulatory enforcement, patient pressure, and commercial incentives. Facilitators included professional identity, peer support, awareness of AMR consequences, and openness to training. These findings highlight the importance of multifaceted, context-specific interventions that address individual behaviour, regulatory structures, and health system enablers. Theory-informed strategies based on the Behaviour Change Wheel (BCW) may enhance antimicrobial stewardship in Ghanaian community settings and similar LMIC contexts.

## Introduction

Antimicrobial resistance (AMR) continues to pose a growing threat to global public health. In 2019, bacterial AMR was associated with 4.95 million deaths and directly responsible for 1.27 million deaths worldwide, with projections suggesting that, without effective interventions, annual fatalities may rise to 10 million by 2050 ^1–4^. This challenge is particularly acute in low- and middle-income countries (LMICs), where high antibiotic use intersects with constrained health infrastructure and surveillance capacity, contributing significantly to the global AMR burden ^5–8^.

Efforts to address AMR have intensified over the past decade, including the launch of the World Health Organization’s (WHO) Global Action Plan on AMR in 2015 and subsequent national action plans ^9–11^. Key initiatives have included regulatory actions to curb substandard and falsified antibiotics, classification of antibiotics using the AWaRe framework to guide appropriate use, and improved monitoring of antimicrobial consumption and resistance trends ^12–15^. Nonetheless, AMR remains a pressing concern in LMICs—particularly in sub-Saharan Africa—where access to reliable diagnostics, robust surveillance systems, and trained healthcare professionals remains limited ^1,10^.

The rising use of WHO ‘Watch’ antibiotics ^15–17^ and the high prevalence of unnecessary prescribing for self-limiting infections, such as upper respiratory tract infections (URTIs), continue to accelerate resistance ^18–23^. Inadequate awareness of antimicrobial stewardship (AMS) principles among healthcare providers and patients further exacerbates the situation ^5,18,24^. Community pharmacies (CPs) and over-the-counter medicine sellers (OTCMS) play a central role in antibiotic supply chains in LMICs and are frequently implicated in the dispensing of antibiotics without a valid prescription—a widespread and persistent practice across sub-Saharan Africa and other LMICs ^5,19–33^.

This pattern of inappropriate dispensing is driven by a combination of limited access to licensed prescribers, high healthcare costs, long travel distances to health facilities, and the commercial imperatives of drug sellers ^4,24,25^. In some cases, community drug outlets, including unlicensed vendors, contribute to the circulation of substandard or falsified antibiotics, compounding the AMR threat ^5,31,32^. While regulatory efforts to restrict non-prescription sales are essential, they may prove impractical in rural or resource-limited settings where healthcare access is fragmented and cost is a major barrier ^5,24^. High out-of-pocket payments remain a key issue across Africa, often leading patients to purchase incomplete courses of antibiotics or rely on stored or shared medicines from family or friends ^25–27^. Pharmacists can play a critical role in mitigating these risks by advising on appropriate use and the importance of completing treatment courses ^24,34^.

In Ghana, existing legislation—including the Pharmacy Act 1994 (Act 489) and the Health Professions Regulatory Bodies Act 2013 (Act 857)—restricts the sale of prescription-only antibiotics without valid prescriptions, with a few exceptions ^34–39^. However, enforcement remains inconsistent, highlighting the need to better understand the behavioural and systemic barriers impeding appropriate dispensing practices among both CPs and OTCMS ^5,24^. Behavioural and social science frameworks offer promising pathways to address these challenges. They consider the broader structural, psychological, and cultural factors shaping dispensing behaviours, moving beyond knowledge-based interventions ^40,43^. The UK Medical Research Council (MRC) advocates the use of theory-informed approaches—such as the Capability, Opportunity, Motivation-Behaviour (COM-B) model and the Theoretical Domains Framework (TDF)—to understand behaviour and inform intervention design ^43^.

Building on our previous simulated client study conducted in Ghana ^33^, this study employs the COM-B and TDF frameworks to explore the barriers and facilitators of appropriate antibiotic dispensing by CPs and OTCMS. The insights generated can inform context-appropriate interventions, guide policy development, and support public education efforts aimed at curbing inappropriate antibiotic use and reducing AMR in Ghana and comparable LMIC settings.

## Materials and Methods

### Study design

This study employed a qualitative design using semi-structured interviews to investigate the behavioural and contextual determinants influencing antibiotic dispensing practices among community pharmacists (CPs) and over-the-counter medicine sellers (OTCMS) in Ghana. The interview guide was informed by two established behavioural science frameworks: the Capability, Opportunity, Motivation-Behavior (COM-B) model and the Theoretical Domains Framework (TDF) ^45–47^. The study adhered to the Consolidated Criteria for Reporting Qualitative Research (COREQ) guidelines to ensure methodological rigor and transparency in the conduct and reporting of qualitative research ^48^.

### Study sites

Participants were purposively recruited from CPs and OTCMS located in four major metropolitan areas across Ghana, collectively representing approximately 14% of the national population ^44^. These sites included two cities in the southern zone—Cape Coast (population: 69,894) and Accra (population: 1,848,614); one city in the central belt—Kumasi (population: 2,035,064); and one city in the northern region—Tamale (population: 371,351). This geographic diversity was selected to capture a broad range of socioeconomic and infrastructural contexts influencing antibiotic dispensing behaviour across the country (**Table 1**).

**Table 1.** Population description and characteristics of the study sties Major Strata Region Major City Population Characteristics.

| Major Strata | Region | Major City | Population Characteristics |
| --- | --- | --- | --- |
| **North** | Northern | Tamale | 1. Low population density<br>2. Relies on agriculture for livelihood<br>3. Limited access to infrastructure which affects connectivity, education and healthcare<br>4. Resident less likely to have insurance |
| **Middle belt** | Asante | Kumasi | 1. Urbanized and economically diverse (i.e agriculture, commercial and industrial activities)<br>2. Serve as central economic hub of Ghana<br>3. Part of the population likely to have insurance |
| **South** | Greater Accra | Accra | 1. Capital city of Ghana<br>2. Most dense population area<br>3. Rich and diverse ethnicity and culture<br>4. Developed infrastructure<br>5. Extensive transport networks, employment, and access to services<br>6. Residents more likely to have health insurance |
|  | Central | Cape Coast | 1. Relies on fishing for livelihood<br>2. Mixed socio-economic landscape<br>3. Inequality in access to healthcare<br>4. Residents likely to have health insurance |

### Sampling and recruitment

Eligible participants were staff members working in registered or licensed community pharmacies (CPs) and over-the-counter medicine seller (OTCMS) outlets who had previously participated in a simulated client study reported by Ngyedu et al. (2023) ^33^. The sample included pharmacists, pharmacy technicians, medical counter assistants (MCAs), and OTCMS personnel. Using contact information obtained from the prior study, the research team initiated recruitment via telephone calls. Potential participants were purposively selected based on their previous demonstration of either appropriate or inappropriate antibiotic dispensing behaviours. During the call, each participant received an explanation of the study objectives and procedures and was invited to participate in a follow-up telephone interview. For those who consented, interviews were scheduled at a time convenient for them.

### Sampling strategy

A maximum variation sampling approach was employed to ensure heterogeneity across socioeconomic and geographic contexts. This strategy also aimed to capture a range of dispensing practices by including a balanced sample of outlets identified in the previous study ^33^: nine exhibiting appropriate practices and three demonstrating inappropriate antibiotic dispensing behaviour.

### Data collection

The semi-structured interview guide was developed based on a review of relevant literature and grounded in the COM-B model to comprehensively explore behavioural influences on antibiotic dispensing ^44–47^. The guide comprised two sections: the first assessed participants’ understanding of AMR, while the second examined barriers and facilitators to appropriate antibiotic dispensing. The guide was piloted with five pharmacy staff members between 3-4^th^ June 2021; data from these pilot interviews were excluded from the final analysis.

Interviews were conducted in English between 1^st^ August and 19^th^ September 2022 and continued until thematic saturation was achieved, defined as the point at which no new themes or insights emerged ^49^. All interviews were audio-recorded, transcribed manually, and independently checked for accuracy by the research team.

### Ethics

Ethical approval for the study was obtained from the Cape Coast Teaching Hospital Ethics Research Committee (CCTHERC/EC/2021/009) (**Supplementary File 1**) and the Pharmacy Council of Ghana (PC-20/4/21) (**Supplementary Files 2 and 3**). Verbal consent was obtained from all participants, which was documented by recording the participant’s explicit verbal agreement at the start of each interview, following a full explanation of the study purpose, procedures, risks, and rights. The interviewer confirmed that the participant understood the information and voluntarily agreed to proceed. Each consent was audio-recorded as part of the interview file and securely stored. In line with ethics requirements, verbal consent was witnessed by the trained interviewer, who documented the date, time, and participant code in the study log. Ethical principles were strictly followed throughout data collection and reporting.

### Data analysis

Data were analysed thematically using an inductive approach. Four members of the research team (GA, MW, JA, and AK) participated in the coding and analysis process. GA began by reading the transcripts repeatedly to ensure data immersion. Transcripts were coded line-by-line, with codes generated inductively from the data. These initial codes were reviewed and refined through iterative discussions within the analysis team. Codes were grouped into subthemes, which were then synthesized into three overarching themes.

While initial mapping of themes to the core COM-B components proved informative, it lacked sufficient granularity. To enhance analytical precision, subthemes were first aligned with specific constructs from the Theoretical Domains Framework (TDF) and subsequently mapped onto the COM-B model ^41,45–47^. This stepwise mapping approach allowed for a more theoretically grounded interpretation of the findings. All transcripts were re-examined to ensure fidelity to the original data, and illustrative quotes were selected to exemplify each theme and subtheme.

The analysis team comprised experts in qualitative research, behavioural science, antimicrobial stewardship (AMS), and health systems implementation, ensuring both methodological rigor and contextual relevance.

## RESULTS

### Participant characteristics

Of the 23 individuals approached for participation, 12 consented and completed the interview process. Among them, eight participants were recruited from community pharmacies and four from over-the-counter medicine shops (OTCMS). Of the eight pharmacy-affiliated participants, five were pharmacists, two were pharmacy technicians, and one was a medical counter assistant. The final sample consisted of 10 male and 2 female participants (**Table 2**).

**Table 2.** Characteristics of the study participants.

| **Participants' characteristic** | **Pharmacies** | **Over-the-counter medical shop** | **Total** |
| --- | --- | --- | --- |
| **Sites** |  |  |  |
| Cape Coast | 3 | 0 | **3** |
| Tamale | 2 | 1 | **3** |
| Kumasi | 1 | 3 | **4** |
| Accra | 2 | 0 | **2** |
| Total | **8** | **4** | **12** |
| **Gender** |  |  |  |
| Female | 1 | 2 | **3** |
| Male | 7 | 2 | **9** |
| **Antibiotic dispensing practices** |  |  |  |
| Appropriate | 1 | 2 | **3** |
| Inappropriate | 7 | 2 | **9** |
| Total | **8** | **4** | **12** |
| **Qualification of pharmacy attendants** |  |  |  |
|  | **Pharmacist** | **Pharmacy Technician** | **Medicine Counter Assistants (MCA)** |
| Cape Coast | 3 | 0 | 0 |
| Tamale | 1 | 1 | 0 |
| Kumasi | 0 | 1 | 0 |
| Accra | 1 | 0 | 1 |
| Total | **5** | **2** | **1** |

### Barriers to and facilitators of appropriate antibiotic dispensing

Three overarching themes were identified through thematic analysis and mapped to the COM-B model, as summarized in Table 3. These included: (1) perceived readiness and capability to dispense antibiotics appropriately (capability); (2) the influence of systemic and contextual factors on dispensing practices (opportunity); and (3) the degree of motivation to adhere to professional guidelines (motivation). Each theme is discussed in detail below.

**Table 3.** Summary of Themes, Subthemes, and Theoretical Mapping of Barriers and Facilitators to Appropriate Antibiotic Dispensing Using the COM-B Model and Theoretical Domains Framework.

| Broad themes | Sub-themes | TDF domains | COM-B |
| --- | --- | --- | --- |
| **1. Perceived readiness and capability to dispense appropriately** | 1. Knowledge of AMR and national guidelines<br>2. Skills and training needed to enhance adherence to guidelines | Knowledge, Memory, Attention and Decision Processes<br>Skills | Psychological Capability<br>Physical capability |
| **2. Impact of a wider system in Antibiotic Dispensing Practice** | 1. Importance of peer support and buy-in<br>2. Patient pressure and expectations<br>3. Environmental factors and resources influencing adherence | Social Influences<br>Social influence<br>Environmental Context and Resources | Social Opportunity<br>Social Opportunity<br>Physical opportunity |
| **3. Making following guidance a priority** | 1. Negotiating professional role and identity | Social/Professional Role and Identity<br>Emotions | Reflective Motivation |
|  | 2. Emotional implications of adhering to the guidelines<br>3. Perceived Benefits and consequences of following the guidelines to practitioners, drug outlet, customers, and public health<br>4. Perception of the impact of AMR and how it emerges | Beliefs about consequences<br>Beliefs about consequences | Automatic Motivation<br>Reflective Motivation |
| (Note): COM-B: Capability, Opportunity, Motivation-Behavior (COM-B); TDF: Theoretical Domains Framework |  |  |  |

### Theme 1: Perceived readiness and capability to dispense appropriately

#### Sub-theme: Knowledge of AMR and national dispensing guidelines (psychological capability)

Participants’ understanding of AMR varied significantly by role and setting. Several OTCMS personnel demonstrated limited conceptual knowledge, often misinterpreting resistance as the body becoming immune to antibiotics or a general loss of drug efficacy. One participant described AMR as:

*“The body is immune to something or the immunity in you has broken down, or is fighting against a particular drug so when you drink, it does not work…” (Participant 06, female, OTCMS, Kumasi)*

By contrast, some OTCMS participants acknowledged AMR as a public health concern and referenced national treatment guidelines, expressing willingness to engage in further training to enhance their knowledge. Pharmacy-affiliated staff generally displayed greater awareness and a more accurate understanding of AMR. They described resistance as the failure of antibiotics to treat infections due to bacterial adaptation and linked its emergence to widespread misuse and overuse of antibiotics within communities. As one technician noted:

*“So, it’s the inability of the antibiotic to fight off infection. Well, it’s very serious in the sense that we dispense the antibiotics and it’s not able to fight the infection…” (Participant 02, male, pharmacy technician, Kumasi)*

Additionally, participants highlighted the absence of diagnostic support at the community level as a contributing factor to inappropriate antibiotic use.

*“It’s due to lack of testing, at the pharmacy level. In the community generally we have the use of, indiscriminate use of antimicrobial.” (Participant 01, female, pharmacist, Tamale)*

Importantly, both pharmacy and OTCMS participants were receptive to further education and training on AMR and appropriate dispensing practices, signaling a potential avenue for targeted stewardship interventions.

#### Sub-theme: Skills and training needed to enhance adherence to guidelines (physical capability)

Participants’ self-reported dispensing skills varied, with most pharmacy-based staff expressing confidence in their ability to validate prescriptions in line with national guidelines. However, some acknowledged only partial competence and emphasized the need for further training to enhance their adherence to recommended practices.

*“I am somehow skilled to dispense antibiotic according to the guidelines, and [am] open for training to improve.”*

*(Participant 12, female, MCA, Accra)*

Pharmacy staff generally reported that they routinely attempted to comply with dispensing protocols. However, practical challenges—such as incomplete or unverifiable prescriptions—frequently undermined their efforts to fully adhere to guidelines.

*“I most of the time remember to follow the guidelines, but sometimes the signatures on the prescription are missing.” (Participant 11, male, pharmacist, Accra)*

Interestingly, some OTCMS personnel with prior medical training perceived themselves as competent in prescription validation, despite legal restrictions prohibiting them from dispensing prescription-only medications in their setting.

*“Not my professional role.” (Participant 03, female, OTCMS, Tamale)*

These findings suggest that while pharmacy personnel generally possess the foundational skills required for appropriate antibiotic dispensing, systemic constraints and regulatory ambiguities—particularly for OTCMS—limit consistent application. Structured training programs, alongside strengthened oversight mechanisms, may be necessary to bridge this gap.

#### Theme 2: Impact of a wider system on antibiotic dispensing practice (opportunity) Sub-theme: Importance of peer support and buy-in (social opportunity)

The influence of peer networks on dispensing practices varied by outlet type. Most OTCMS participants reported operating individually or under family-run models, often lacking professionally trained staff. As a result, peer influence was minimal or non-existent. In some cases, untrained family members assumed responsibility in the dispenser’s absence:

*“For him (sibling), when I am around, but when I am going out I leave the store in his care.” (Participant 06, female, OTCMS, Kumasi)*

In contrast, some pharmacists reported a more collaborative work environment where colleagues encouraged adherence to national dispensing guidelines:

*“They know the guidelines are there to help the patients, so they support it.” (Participant 01, female, pharmacist, Tamale)*

However, other participants highlighted that the presence of non-pharmacist staff could create tension or resistance to guideline adherence:

*“At the pharmacy, maybe the pharmacist [complies] but the rest of the staff… they are all not pharmacy inclined so they wouldn’t feel pleased.” (Participant 10, male, pharmacist, Cape Coast)*

Some pharmacy technicians noted that their dispensing decisions were driven by personal professional standards, irrespective of peer input:

*“We all have individual mindsets… So professionally, you have to follow what your profession is telling you… I am good to go since I am doing the right part.” (Participant 09, male, pharmacy technician, Tamale)*

### Sub-theme: Patient pressure and expectations (social opportunity)

Many participants reported that patient demands were a major barrier to appropriate dispensing. Clients often insisted on obtaining antibiotics without prescriptions, particularly in long-standing customer relationships:

*“Customers make it difficult by insisting on you giving them a prescription drug… because you are a community pharmacy, we’ve been with them for long. So, they will be like ‘we are buying from you and you don’t want to sell it.” (Participant 12, female, MCA, Accra)*

Compounding this pressure, healthcare workers occasionally sought to bypass prescription *requirements based on their professional status, challenging the authority of pharmacists:*

*“A health worker… some are not, excuse me to say, ‘being reasonable.’ I understand that you have some knowledge and education, but what makes me a pharmacist is being a custodian of medication… most of these health workers… come with [the] excuse of ‘I am a health worker.’” (Participant 01, female, pharmacist, Tamale)*

### Sub-theme: Environmental factors and resource constraints (physical opportunity)

Participants widely agreed that infrastructure and tools—such as digital systems or prescription verification logs—could support compliance with antibiotic dispensing guidelines. However, most reported the absence of such resources, including dedicated record-keeping systems:

*“We don’t have an antibiotic book for record-keeping.” (General sentiment across participants)*

Some highlighted that access to computers and sufficient time enabled better patient counseling and adherence monitoring. Yet, profitability pressures and patient poverty frequently undermined guideline adherence:

*“Here lies a case where someone brings a prescription which would serve [generate] much money for the company, but you doubt it… So you call to validate, and the patient says, ‘If you won’t sell, let me go to another outlet.” (Participant 11, male, pharmacist, Accra)*

Financial constraints were also reported as a major barrier to appropriate prescribing and dispensing, particularly in resource-limited areas where patients struggled to afford laboratory testing or physician consultations:

*“A lot of patients, especially in this part of the country where the standard of living is pretty low, have just enough money to buy the drug but not to run a lab… If you run the lab, you are not going to get the drug and vice versa.” (Participant 01, female, pharmacist, Tamale)*

These findings underscore the influence of broader health system limitations—such as poverty, business models, and lack of infrastructure—on dispensing behaviors in both formal and informal settings.

## Theme 3: Committing to Following the Guidance - Motivation

### Sub-theme: Negotiating professional role and identity (reflective motivation)

Pharmacists consistently viewed adherence to national antibiotic dispensing guidelines as integral to their professional identity and ethical responsibility. They recognized the importance of dispensing only with valid prescriptions as part of their formal training:

*“Part of professional role to adhere to the guidelines.” (Participant 08, male, pharmacist, Cape Coast)*

In contrast, pharmacy technicians and medical counter assistants (MCAs) often viewed the enforcement of appropriate dispensing practices as a shared responsibility, particularly emphasizing the role of the Pharmacy Council in ensuring regulatory compliance:

*“I think when the regulators do their work well, always making sure a pharmacy shop has a pharmacist, and chemical shops don’t have to stock these drugs…” (Participant 02, male, pharmacy technician, Kumasi)*

While this suggests some diffusion of responsibility, participants still expressed a collective commitment to mitigating antimicrobial resistance (AMR), acknowledging that the consequences of inappropriate antibiotic use affect both the profession and public health. However, some participants indicated that despite understanding their role, they had not fully internalized personal accountability for ensuring compliance with dispensing guidelines.

### Sub-theme: Emotional implications of adhering to the guidelines (automatic motivation)

Most participants reported that emotional appeals from customers did not significantly alter their dispensing decisions, although they acknowledged the emotional burden of refusing antibiotics—especially in cases involving longstanding customers or individuals expressing distress:

“*People come in with all sorts of emotions… just to make you feel in a way bad, to give it to them, to have pity on them and all that.” (Participant 11, male, pharmacist, Accra)*

Some participants experienced regret or guilt when they were unable to assist regular customers or when adherence to the guidelines conflicted with customer expectations:

*“I didn’t feel right because she was a customer and she thought I could have done her that favor.” (Participant 12, female, MCA, Accra)*

Among OTCMS participants, adherence to the guidelines was further complicated by concerns over lost income. The commercial nature of their practice heightened emotional discomfort when declining antibiotic sales:

*“Most of the time it’s money that we think about… but as for what happens after, people do not care.” (Participant 04, female, OTCMS, Kumasi)*

### Sub-theme: Perceived benefits and consequences of following the guidelines (reflective motivation)

Several pharmacists emphasized the long-term professional benefits of guideline adherence, including enhanced trust, credibility, and patient satisfaction:

*“I will gain the trust of my clients and the satisfaction of knowing I am taking care of someone to the best of my ability.” (Participant 01, female, pharmacist, Tamale)*

Participants widely believed that consistent compliance could bolster the reputation of their facility and attract more customers:

*“The pharmacy will be noted to be serving good drugs… knowing very well that we adhere to the rules and regulations.” (Participant 07, male, pharmacist, Cape Coast)*

OTCMS participants also noted that adherence reduced fear of regulatory inspections and contributed to a more secure and legitimate practice:

*“The benefit will be that I will have no fear that maybe some people will come for inspection… and probably pack [the drugs] away.” (Participant 05, male, OTCMS, Kumasi)*

Although many acknowledged that compliance could improve patient safety and reduce long-term costs, some were concerned about losing customers who might turn to less regulated sources if denied antibiotics:

*“For my customers… when they reduce and your sales drop… you will lose customers also.” (Participant 06, female, OTCMS, Kumasi)*

Nevertheless, others remained optimistic that consistent enforcement could gradually shift customer expectations toward more responsible antibiotic use.

### Sub-theme: Perception of the impact of AMR and how it emerges (reflective motivation)

All pharmacists expressed serious concern about AMR, citing direct experiences with treatment failure and resistance patterns observed in hospital laboratories. These clinical encounters reinforced their motivation to preserve the effectiveness of current antibiotics:

*“Most of these samples taken are resistant to almost all our antibacterials… So the question is what is next? We have to make use of the ones we have currently or… we will not be able to curb simple bacterial infections with our common antimicrobials.” (Participant 01, female, pharmacist, Tamale)*

Participants broadly recognized that inappropriate antibiotic use—especially unsupervised purchasing and incomplete courses—drives resistance in the community, leading to reduced treatment effectiveness, higher healthcare costs, and avoidable morbidity. They viewed adherence to dispensing guidelines as a key strategy to prevent AMR-related harm:

*“Benefits for public health are definitely going to contribute to the decline in resistance… antibiotics that were commonly used will be effective for treatment… it will reduce the cost of treatment, reduce hospital stays, and even fatality.” (Participant 11, male, pharmacist, Accra)*

## Discussion

### Key findings

To our knowledge, this is the first study in Ghana and possibly Africa to apply behavioural science frameworks—specifically the COM-B model ^41^—to explore the determinants of antibiotic dispensing behaviours among both CPs and OTCMS. Through thematic analysis, we identified three interrelated themes mapped to the COM-B components: (1) perceived readiness and capability to dispense appropriately (capability), (2) the influence of systemic and contextual factors (opportunity), and (3) professional motivation to comply with guidelines (motivation) ^44–47^.

Marked variability was observed in participants’ physical and psychological capabilities, particularly in their knowledge of AMR, familiarity with national dispensing guidelines, and ability to validate prescriptions. Notably, knowledge gaps were more pronounced among OTCMS, a finding consistent with previous studies highlighting the role of inadequate training and weak regulatory oversight in driving inappropriate antibiotic access across Africa—including for WHO Watch list antibiotics ^5,24,50,51^. These findings underscore the need for robust, ongoing educational and AMS interventions targeting all community dispensers.

While many participants expressed confidence in validating prescriptions and understanding national protocols, practical adherence was hindered by factors such as missing prescriber signatures and limited enforcement of regulatory policies. Similar barriers have been documented in the literature, where inconsistent enforcement remains a persistent issue in LMIC settings ^5,24^. Participants with hospital experience noted greater compliance in institutional settings due to structured prescription validation systems, including electronic platforms—a finding supported by studies demonstrating the positive impact of digital tools on prescribing behaviours ^52–54^. In contrast, the absence of such systems in community settings revealed a critical gap between self-reported competence and actual practice, particularly among OTCMS who reported dispensing prescription-only antibiotics despite legal prohibitions. This highlights regulatory lapses and role ambiguity that warrant urgent attention.

Dispensing behaviour was also shaped by social and environmental opportunities. External pressures such as customer demands, peer influence, and business-driven incentives contributed to deviations from best practice. These drivers echo findings from Ghana, Nigeria, and other LMICs, where informal dispensing is heavily influenced by sociocultural norms and economic imperatives rather than clinical guidelines ^28,32,34,39,55^.

Affordability emerged as a major barrier, with many patients unable to afford diagnostic testing or physician consultations. In such contexts, drug outlets faced commercial pressures to meet patient demands, leading to inappropriate antibiotic provision. These findings align with prior reports on the impact of high patient co-payments and economic constraints on antimicrobial use in Africa ^5,24^. Application of the COM-B model suggests that bundled interventions integrating education, persuasion, and financial incentives may be necessary to promote behaviour change ^40–42^. Current supervisory models in Ghana, which rely largely on in-person inspection, may be insufficient to drive sustained practice improvements.

A disconnect was observed between participants’ professional identity and their actual dispensing behaviour. While pharmacists acknowledged that dispensing antibiotics without a prescription contravenes national laws, compliance was inconsistent—mirroring prior reports of widespread non-compliance across Ghana and other LMICs ^15,19,20,31–34,39,55^. Many participants, particularly non-pharmacist staff, perceived guideline enforcement as the sole responsibility of the Pharmacy Council, which may undermine intrinsic motivation. Strengthening personal accountability through targeted training and institutional support could help address this gap.

Despite these challenges, many participants recognized the long-term benefits of adherence—including enhanced patient trust, professional credibility, and public health gains. However, emotional appeals from regular clients, or fear of financial loss, often led to deviations from appropriate practice—especially among OTCMS. These relational dynamics have also been documented in other LMIC settings ^26,31,32,34,56–59^. Notably, participants demonstrated awareness of AMR’s clinical and economic impact, with some pharmacists drawing on hospital experience to describe treatment failures due to multidrug-resistant infections. This awareness represents a potential foundation for behaviour change interventions.

### Strengths and limitations

This study is among the few in sub-Saharan Africa to apply a behavioural science lens to antibiotic dispensing practices in the community. The inclusion of diverse professional roles and geographic regions—encompassing both rural and urban contexts—enhances the generalizability of findings within Ghana. Methodological rigor was upheld through adherence to the COREQ guidelines and use of multiple independent coders to ensure analytical reliability.

However, the study has limitations. The relatively small number of OTCMS participants may have restricted the range of perspectives from this group. This likely reflects hesitancy to participate due to concerns over disclosing potentially non-compliant practices. Future research should incorporate patient perspectives and build on existing studies among prescribers to provide a more holistic view of the systemic drivers of antibiotic misuse.

## Conclusion

Improving antibiotic dispensing behaviour in Ghana requires interventions that address all three domains of the COM-B model. Enhancing knowledge and dispensing skills (capability) must be complemented by supportive systems and regulatory reforms (opportunity), including standardized prescription formats, accessible validation mechanisms, and strengthened supervisory structures. Simultaneously, efforts to reinforce professional identity and individual accountability (motivation) are critical for sustainable change. Without such comprehensive, contextually tailored strategies, the disconnect between policy and practice will persist—limiting national and global efforts to curb antimicrobial resistance. By applying the COM-B framework, this study offers a theory-informed foundation for designing multifaceted interventions to reduce inappropriate antibiotic dispensing in Ghana and similar LMIC settings.

## Data Availability

The data underlying this study consist of anonymized interview transcripts. Due to ethical restrictions related to participant confidentiality and the sensitivity of professional and legal disclosures regarding antibiotic dispensing, the full transcripts cannot be made publicly available. However, relevant excerpts supporting the findings are included within the manuscript. Researchers who meet the criteria for access to confidential data may request access to de-identified data by contacting the Cape Coast Teaching Hospital Ethics Research Committee (CCTHERC/EC/2021/009) at or the corresponding author at

## Acknowledgement

We thank Mr. Samuel Osae and Mr. Jonathan Amartey, both clinical psychologists at the Cape Coast Teaching Hospital who conducted the interviews for their contributions to the study. Thank you to Dave David for transcribing the interviews. Special thanks to Renee Frances Wassick and the family for all the support throughout the work. We also wish to thank the Pharmacy Council of Ghana for providing the necessary support to conduct this study. This work has not been previously presented as an abstract or at a conference or similar forum. George Akafity received guidance through Professor Jordi Rello through the ESCMID mentorship program.

## Author Contribution Statement

Study conception and design: all authors; data collection and management: GA (George Akafity); data analysis and interpretation: GA (George Akafity), MWat (Marta Wanat), and JA (Joseph Acolatse); manuscript writing and drafting: GA (George Akafity), BG (Brian Godman), MWat (Marta Wanat), AK (Amanj Kurdi), JA (Joseph Acolatse); manuscript reviewing and revising as well as final approval: all authors. All authors have substantially contributed to the conception and design of the article and interpreting the relevant literature and have been involved in writing the article or revised it for intellectual content.

